# SARS-CoV-2 transmission in cancer patients of a tertiary hospital in Wuhan

**DOI:** 10.1101/2020.02.22.20025320

**Authors:** Jing Yu, Wen Ouyang, Melvin L.K. Chua, Conghua Xie

## Abstract

In December 2019, an outbreak of atypical pneumonia known as 2019 novel coronavirus disease (COVID-19) occurred in Wuhan, China. This new type of pneumonia is characterized by rapid human-to-human transmission. Among the different disease types, cancer patients are often recalled to the hospital for treatment and disease surveillance, and the majority of cancer treatments such as chemotherapy and radiotherapy are immunosuppressive. This prompts us to consider if cancer patients were at an elevated risk of SARS-CoV-2 infection.

A total of 1,524 cancer patients who were managed at our tertiary cancer institution – Zhongnan hospital of Wuhan University were reviewed during the period of Dec 30, 2019 to Feb 17, 2020. It was found that cancer patients had an estimated 2-fold increased risk of COVID-19 than the general population. We identified twelve patients who were infected with SARS-CoV-2, with two recorded deaths (16.7%), albeit one patient passed away from a COVID-19 unrelated cause. Interestingly, only five of these patients were ongoing treatment at the time of contracting the virus, suggesting that hospital visitation was the likely factor contributing to the elevated incidence in cancer patients. Moreover, we also observed that the incidence of severe COVID-19 was not higher than in the general population. Consequently, for cancer patients who require treatment, proper isolation protocols must be in place to mitigate the risk of SARS-CoV-2 infection.

---

In December 2019, an outbreak of atypical pneumonia known as 2019 novel coronavirus disease (COVID-19) occurred in Wuhan, Hubei, China. This new type of pneumonia is linked to the severe adult respiratory syndrome coronavirus 2 (SARS-CoV-2), and is characterized by rapid human-to-human transmission, patients would develop progressive respiratory symptoms upon infection.^1^ Droplet transmission appears to be the main route of transmission for SARS-CoV-2.^2^ According to a recent report of 138 hospitalized patients from a single institution (Zhongnan Hospital of Wuhan University), it was indicated that hospital-acquired transmission accounts for 41.3% of these admitted patients (29% were hospital staff and 12.3% were in-house patients), thereby highlighting that the hospital environment may be a hotspot for viral spread.^3^ Among the different disease types, cancer patients are often recalled to the hospital for treatment and disease surveillance, and therefore, they may be at an elevated risk of contracting SARS-CoV-2. Moreover, the majority of cancer treatments such as chemotherapy and radiotherapy are immunosuppressive. Here, we report the incidence and outcomes of SARS-CoV-2 infection in cancer patients who were managed at a tertiary cancer institution in Wuhan.

We collected the data of all patients from Dec 30, 2019 to Feb 17, 2020 (data cut-off date) who were admitted to the Department of Radiation and Medical Oncology, Zhongnan Hospital of Wuhan University. In total, 1,524 cancer patients were included in our study; the three commonest cancer diagnoses were gastrointestinal (N = 394, 35.9%), thoracic (N = 326, 21.4%), and head and neck cancers (N = 204, 13.4%). Among them, 12 patients were diagnosed with COVID-19 pneumonia (four by the real-time reverse transcription polymerase chain reaction assay for SARS-CoV-2 and eight by the clinical criteria of fever and radiological computed tomography changes; **Table 1**). We estimated that the infection rate of SARS-CoV-2 in cancer patients from our single institution at 0.79% (95% CI = 0.3–1.2). This was higher than the cumulative incidence of all diagnosed COVID-19 cases that was reported in the city of Wuhan over the same time-period (0.37%, 41,152/11,081,000 cases, data cutoff on Feb 17, 2020).

**Table 1.** Clinical characteristics and outcomes of cancer patients with severe adult respiratory syndrome coronavirus 2 (SARS-CoV-2) infection.

| Case | Sex | Age | P<br>S | Cancer<br>diagnosis | Phase of cancer treatment | Date of<br>COVID-19<br>diagnosis | Date of first<br>onset of<br>symptoms | Fever | Dyspnea | Cough | CT<br>diagnosis | SARS-COV-2 RT-PCR<br>assay | Severe events<br>due to<br>COVID-19 | Hospitalization<br>status | Survival<br>status |
| --- | --- | --- | --- | --- | --- | --- | --- | --- | --- | --- | --- | --- | --- | --- | --- |
| 1 | Male | 57 | 1 | NSCLC | Concurrent EGFR TKI (osimertinib [80 mg/day]) with concurrent thoracic radiotherapy ((27 Gy in 9 fr)) | 2020/1/26 | 2020/1/18 | Yes | No | No | Positive | Positive | No | Discharged | Alive |
| 2 | Male | 69 | 1 | NSCLC | W5 of chemoradiotherapy (Chemotherapy regime: pemetrexed [500mg/m<sup>2</sup>], cisplatin [75mg/m<sup>2</sup>] q3W; Radiotherapy 54 Gy in 27 fr) | 2020/1/18 | 2020/1/17 | Yes | Yes | Yea | Positive | Negative | No | Admitted | Alive |
| 3 | Femal<br>e | 78 | 3 | NSCLC | Admitted for pleural effusion; best supportive care | 2020/1/17 | 2020/1/17 | Yes | Yes | Yes | Positive | Negative | No | Discharged | Alive |
| 4 | Male | 60 | 1 | NSCLC | Follow-up at 2 years post-chemoradiotherapy; no evidence of disease relapse | 2020/2/14 | 2020/2/14 | Yes | Yes | Yes | Positive | Negative | Yes | - | Dead |
| 5 | Male | 69 | 1 | NSCLC | Post-first cycle (docetaxel, [75mg/m<sup>2</sup>], cisplatin [75mg/m<sup>2</sup>], sintilimab, [200mg]) | 2020/1/22 | 2020/1/22 | Yes | No | No | Positive | Negative | No | Discharged | Alive |
| 6 | Male | 66 | 1 | NSCLC | Post-third cycle (pemetrexed [500mg/m<sup>2</sup>], carboplatin [AUC5], pembrolizumab [200mg]) | 2020/1/22 | 2020/1/22 | Yes | No | No | Positive | Negative | No | Admitted | Alive |
| 7 | Male | 65 | 1 | NSCLC | Surgery (April 23th, 2019), Post-four cycles adjuvant chemotherapy (docetaxel [75mg/m<sup>2</sup>], cisplatin [75mg/m<sup>2</sup>]) | 2020/1/20 | 2020/1/20 | Yes | No | No | Positive | Negative | No | Discharged | Alive |
| 8 | Male | 76 | 2 | Rectal cancer | Best supportive care | 2020/1/22 | 2020/1/22 | Yes | No | No | Positive | Negative | No | Admitted | Alive |
| 9 | Male | 57 | 1 | Colon cancer | Newly diagnosed; treatment yet to commence | 2020/2/13 | 2020/1/21 | Yes | No | No | Negative | Positive | No | Admitted | Alive |
| 10 | Male | 67 | 3 | Pancreatic cancer | Best supportive care | 2020/1/22 | 2020/1/22 | Yes | No | No | Positive | Negative | Yes | - | Dead |
| 11 | Femal<br>e | 48 | 1 | Breast cancer | Adjuvant radiotherapy (42 Gy in 21 fr) | 2020/1/26 | 2020/1/24 | Yes | No | No | Positive | Positive | No | Admitted | Alive |
| 12 | Male | 77 | 2 | Urothelial cancer | Best supportive care | 2020/2/17 | 2020/2/11 | Yes | No | No | Not performed | Positive | No | Admitted | Alive |

Abbreviations: PS, performance status; CT, computed tomography; RT-PCR, real-time quantitative polymerase chain reaction; COVID-19, 2019 coronavirus disease; SARS-CoV-2, severe acute respiratory syndrome coronavirus 2; NSCLC, non-small cell lung cancer.

Clinical details on the cancer history and COVID-19 pneumonia are summarized in Table 1. Among the SARS-CoV-2 infected patients, 58.3% (7 of 12) were non-small cell lung carcinoma patients. Five (41.7%) were on-going treatment with either chemotherapy +/- immunotherapy (N = 3) or radiotherapy (N = 2; one patient was undergoing concurrent chemoradiotherapy). Two patients (0.17%) developed severe adult respiratory distress syndrome (SARS), but none of the on-treatment patients required intensive-level care. Four patients (33.3%) have been discharged, and two deaths were recorded; one patient died from a COVID-19 unrelated cause.

It is hypothesized that cancer patients may be susceptible to an infection during a viral epidemic due to their compromised immune status, which may be induced by the treatment of the cancer. ^4^ Based on our single institution data, we observed that cancer patients from the epicenter of a viral epidemic indeed harbor a higher risk of SARS-CoV-2 infection (OR = 2.31, 95% CI = 1.89-3.02) when compared to the general community. Nonetheless, only less than half of these infected patients were undergoing active treatment for their cancers. It is arguable that a third of the patients were terminally admitted for best supportive measures, and these patients are likely immunosuppressed. Regardless, our findings do imply that hospital admission and recurrent hospital visits are potential risks for SARS-CoV-2 infection, and these factors may account for the increased incidence among cancer patients. We therefore propose that aggressive measures are undertaken to reduce frequency of hospital visits during a viral epidemic going forward. For cancer patients who require treatment, proper isolation protocols must be in place to mitigate the risk of SARS-CoV-2 infection.

## Data Availability

Clinical details on the cancer history and COVID-19 pneumonia are summarized in Table 1. The further details of the current study are available from the corresponding author upon reasonable request.

---

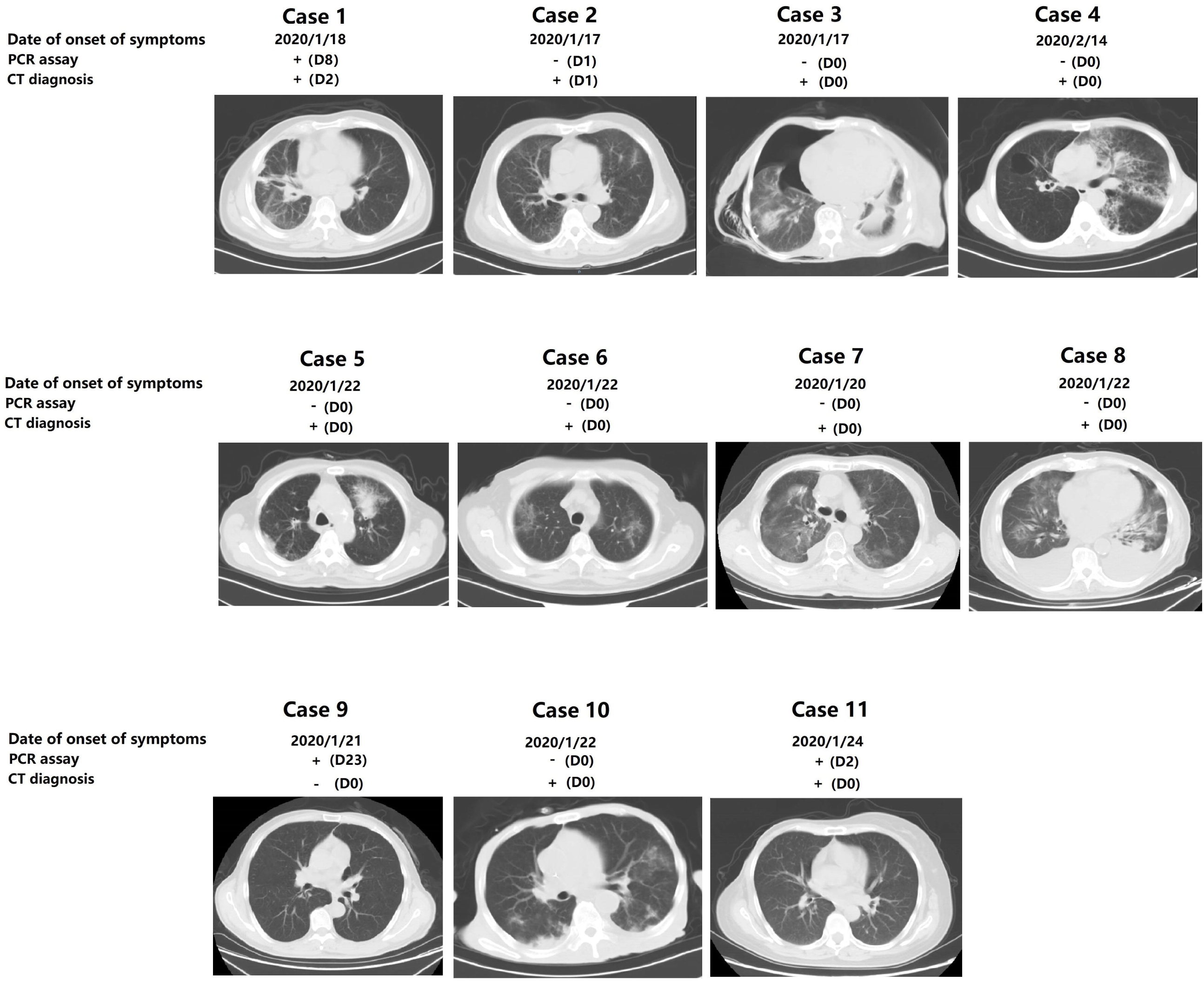

## Notes

### Competing Interest Statement

The authors have declared no competing interest.

### Clinical Trial

This study was retrospective.

### Funding Statement

There is no funding for the current study.

## REFERENCES

1. Chen N, Zhou M, Dong X, et al. Epidemiological and clinical characteristics of 99 cases of 2019 novel coronavirus pneumonia in Wuhan, China: a descriptive study. Lancet (London, England) 2020.

2. Expert consensus for bronchoscopy during the epidemic of 2019 Novel Coronavirus infection (Trial version). Chinese journal of tuberculosis and respiratory diseases 2020;43:E006.

3. Wang D, Hu B, Hu C, et al. Clinical Characteristics of 138 Hospitalized Patients With 2019 Novel Coronavirus-Infected Pneumonia in Wuhan, China. Jama 2020.

4. Kamboj M, Sepkowitz KA. Nosocomial infections in patients with cancer. The Lancet Oncology 2009;10:589–97.

